# Association of age-friendly communities with health and well-being among older adults: an ecological and multilevel analysis from the Japan Gerontological Evaluation Study

**DOI:** 10.1101/2024.06.21.24309218

**Authors:** Taiji Noguchi, Satoko Fujihara, Kazushige Ide, Seungwon Jeong, Tami Saito, Katsunori Kondo, Toshiyuki Ojima

## Abstract

We examined the association of age-friendly communities with health and well-being among older adults in Japan. Ecological and multilevel analyses of 71,824 older adults across 145 communities revealed that the community’s age-friendliness consistently showed associations with health and well-being. Age-friendly physical environments (accessibility to barrier-free outdoor spaces, buildings, and transportation resources) exhibited an inverse association with functional health deficits. Social engagement and communication (participation in community groups, volunteer engagement, and information use) were inversely associated with depressive symptoms. Social inclusion and dementia-friendliness (respect and inclusion for older adults and positive attitudes toward people with dementia) were positively associated with happiness. The community’s age-friendliness reflected well the multiple aspects of older adults’ health and well-being.

## Introduction

In many countries, the population of older adults is experiencing the most rapid growth. It is projected that the number of individuals aged 65 years or above will surge from 771 million in 2022 to 1.6 billion by 2050, with their global proportion nearly rising from 9.7% in 2022 to 16.4% in 2050 (United Nations Department of Economic and Social Affairs, 2022). The evolving needs of older residents necessitate supportive living conditions to respond to the physical, mental, and social changes experienced as a result of biological aging (World Health Organization, 2020). Therefore, it is imperative to address both the social and physical aspects of the community environments to effectively cater to the needs and preferences of older adults and enhance their health and well-being.

To achieve healthy aging, living longer with health and well-being, the promotion of “age-friendly communities” (AFC) is becoming active worldwide (World Health Organization, 2007, 2015, 2023). The AFC is regarded as an inclusive and accessible community environment that optimizes opportunities for health, participation, and safety for all individuals, to ensure the quality of life and dignity as people age (World Health Organization, 2015). World Health Organization (WHO) has identified a total of eight topic areas that comprehensively cover the age-friendliness of the communities, including the features of the structures, environments, services, and policies of communities that reflect the determinants of active aging (World Health Organization, 2007, 2015, 2023). Establishing the region-appropriate AFC scales and evaluating the community’s age-friendliness in line with this framework helps promote a common understanding of AFC among stakeholders and encourages monitoring and action on AFC-related policies and practices (World Health Organization, 2015).

However, there are at least three challenges left for AFC studies. First, as for the AFC assessment, the measurement on a community or neighborhood basis was not sufficient. Although, several previous studies have attempted to assess the age-friendliness of the communities based on these topic areas (Dikken et al., 2020; Garner and Holland, 2020; Kim et al., 2022; Lehning et al., 2014; Özer et al., 2023; Xie, 2018), most existing scales are limited to those that measure individual-level perceptions and attitudes. Given that the AFC framework is a community or neighborhood feature and characteristics for age-friendliness, providing community-level AFC assessments beyond the individual level would be of considerable meaning for community assessment and policy-making for healthy aging. In the United States (Lynott et al., 2018), South Korea (Park and Lee, 2017), and China (Xu et al., 2022), measurement tools for age-friendliness at the community or neighborhood levels have been developed; nevertheless, there is still a small scientific accumulation of these measures internationally.

Second, empirical evidence linking AFC to outcomes in older adults is scarce. The WHO provides the health and well-being outcomes of the residents as impact indicators of the AFC, which constitute long-term changes to be achieved as a result of the promotion and improvement of the AFC, and can be well reflected in the realization of age-friendly physical and social community environments (World Health Organization, 2015). Several prior studies link AFC features to superior outcomes. The activeness of older adults’ social participation in the community correlates well with community-level higher levels of happiness (Ide et al., 2022). Those who rated communities they lived in as more age-friendly reported better health (Fennis et al., 2015; Lehning et al., 2014; Yu et al., 2019) and well-being (Au et al., 2020; Gibney et al., 2020; Xie, 2018). However, it highlights the necessity for multilevel analysis, as well as ecological or individual analysis, to explore the links between the community characteristics of age-friendliness and health and well-being. Although ecological studies of AFC have yielded useful insights because of helping community assessment or monitoring on the areas, a proper examination of community-level characteristics as a collective or contextual influence on older adults’ outcomes requires multilevel analysis (Kawachi et al., 2008). Multilevel analysis, which assumes nested structures of individuals (micro levels) within communities or neighborhoods (macro levels), can address the contextual effects of community features on individual outcomes (Diez Roux, 2015). This means that multilevel approaches help explore to enhance older adults’ health and well-being through developing age-friendly communities. Some studies using multilevel analysis have reported that a community’s age-friendliness is linked to individual perceived health (Choi, 2020), functional health (Choi, 2020), psychological and mental health (Park and Lee, 2017), and frailty (Xu et al., 2022). However, the evidence remains sparse and it is required to determine whether AFC would be well reflective of the impact indicators of health and well-being using the multilevel framework.

Third, it is not addressed in the evaluation of communities’ age-friendliness including the aspects of dementia-friendliness. Given the growing number of individuals with dementia living in the community, dementia-friendly communities (DFC) should be increasingly considered (Altzeheimer’s Disease International, 2016; Hung et al., 2021). DFC is the involvement of people living with dementia in all aspects of their organization and operations, an approach of promoting their social participation and recognizing their human rights through removing socially imposed barriers and focusing on enablement (Altzeheimer’s Disease International, 2016; United Nations Department of Economic and Social Affairs, 2006). Age-friendly and dementia-friendly approaches share similarities and some fundamental objectives, for instance, aiming to help older adults remain independent and in the community as long as possible through creative, supportive, and enabling environments (AARP International Affairs, 2016; Rahman and Swaffer, 2018). However, some have pointed out that AFC is not necessarily dementia-friendly (AARP International Affairs, 2016; Dementia Australia; Rahman and Swaffer, 2018). Age-friendliness strategies encompass a holistic view of older adults without identifying people solely through the disease-specific lens, while dementia-friendliness is more targeted and disease-specific. DFC initiatives span from social environments related to social respect and support for individuals living with dementia to physical environments such as dementia-friendly structures and designs such as transportation and buildings (Altzeheimer’s Disease International, 2016; Diaz et al., 2022). In particular, the beliefs and attitudes of residents in the community regarding the social inclusion of individuals with dementia and their families are key elements of the DFC as these can pose social exclusion and barriers to their social engagement (Diaz et al., 2022). Age-friendliness strategies can provide benefits to older adults or individuals with disabilities more generally, but they do not necessarily cover dementia-friendly actions and design features that address a particular set of needs. Considering the suggestions for implementing dementia-friendliness in the broader context of the AFC (World Health Organization, 2021), it is crucial to evaluate the AFC while incorporating indicators that consider the dementia-friendly aspects of communities.

Recently, an AFC assessment scale, encompassing dementia-friendly elements, was developed in Japan, one of the world’s fastest-aging nations (Fujihara et al., 2024). This scale offers community-level assessment by aggregating individual responses by community unit, which covers the almost core areas of the AFC outlined by the WHO, and incorporates elements of a dementia-friendly social environment, such as positive feelings and attitudes towards the social participation of individuals with dementia and supportive awareness of family members of individuals with dementia (Fujihara et al., 2024). The scale is composed of three factorial groups (“age-friendly physical environments,” characterized by accessibility to barrier-free outdoor spaces and buildings and transportation, “social engagement and communication,” characterized by social activities in the community group, volunteer engagement, and information communication, and “social inclusion and dementia-friendliness,” characterized by respect and inclusion of older adults and dementia-friendly briefs and attitudes) and has demonstrated validity and reliability (Fujihara et al., 2024). However, it is unclear whether community-level age-friendliness assessed using this scale would link to the health and well-being of the older population in the community.

Accordingly, this study aimed to elucidate the associations of AFC with health and well-being among older adults, using a community-level assessment scale of the AFC including the elements of dementia-friendliness. We approached compositional and contextual effects on the association between community-level age-friendliness and outcomes, through multilevel analysis as well as ecological one.

## Methods

### Study participants

This cross-sectional study of the ecological and multilevel analysis design used data from the 2016 wave of the Japan Gerontological Evaluation Study (JAGES), an ongoing cohort study among older adults aged 65 years and above in Japan, without receiving public long-term care insurance benefits (Kondo et al., 2018).

Supplementary Figure 1 shows the sample selection flow. Self-administered questionnaires were distributed through mail to older adult residents in 39 municipalities; the municipalities were not randomly selected but covered a wide range of characteristics in terms of regions and population sizes in Japan. Random sampling methods were used in 17 large municipalities, while all eligible residents were sampled in 22 small municipalities. Several items related to the AFC scale were randomly assigned to survey questionnaire modules in one-eight of all participants. Among the total of 270,661 residents invited to participate, 196,438 returned the questionnaires (response rate = 70.2%). We excluded 16,417 respondents whose age and/or sex could not be confirmed or who received public long-term care insurance benefits (valid response rata = 64.4%). Subsequently, we defined the school districts as community units and excluded 87,511 individuals in 849 community areas with less than 30 respondents regarding all AFC survey modules randomly assigned to part of the participants in order to avoid non-precision due to the small sample size; 9,899 respondents with unknown areas of residence were also excluded. Additionally, those with care needs in daily life (n = 5,175) and with missing information on the item (n = 5,612) were excluded. Thus, we derived data from 71,824 individuals covering 145 communities in the final analysis (the mean number of observations per community = 495.3; min–max = 216–1,859).

This study was reviewed and approved by the ethics committees on Human Subjects at the National Center for Geriatrics and Gerontology (No. 992) and Chiba University (No. 2493). The mailed questionnaire was accompanied by a study explanation; participants who returned the completed questionnaire were considered to have provided informed consent. All our procedures conformed to the Declaration of Helsinki.

### Exposures: age-friendly community scale

Based on a prior study (Fujihara et al., 2024), we assessed communities using the AFC scale. Individual-level responses on the AFC scale items were aggregated by school districts. We selected school districts as community units where older adults can easily travel by foot or bicycle and are a reasonable unit for considering local public health activities (Saito et al., 2017). This scale consists of three domains with 17 items: (i) age-friendly physical environments, (ii) social engagement and communication, and (iii) social inclusion and dementia-friendliness. Supplementary Table 1 presents the details of the scale items and Supplementary Table 2 shows their factorial structure. The domain of age-friendly physical environments consists of five items characterized by accessibility to barrier-free outdoor spaces and buildings and transportation (scores range from 0–500 points; a higher score indicates a more age-friendly physical environment). The domain of social engagement and communication includes five items characterized by older adults’ social participation, volunteer employment, and information communication (score ranges from 0–500 points; a higher score indicates more active engagement and communication among older adults). Finally, the domain of social inclusion and dementia-friendliness comprised seven items characterized by the elements of respect and inclusion for older adults as well as dementia-friendly feelings and attitudes (scores range from 0–700 points; a higher score indicates more inclusive and dementia-friendliness). This scale was confirmed for factorial validity and retest reliability, with each domain having a Cronbach’s alpha coefficient ≥ 0.78 (Fujihara et al., 2024).

### Outcomes: health and well-being

As the impact indicators of AFC (World Health Organization, 2015), four outcomes of health and well-being were assessed: self-reported health, happiness, depressive symptoms, and functional health. These are valid predictors of mortality and disability, regardless of other medical, behavioral, or psychosocial factors (Diener and Chan, 2011; Fujiwara et al., 2003; Nagata et al., 2023; Watanabe et al., 2023; Wuorela et al., 2020). Self-reported health was assessed using the question, “How do you feel about your current health status?” (possible answers: “excellent,” “good,” “fair,” or “poor”), which was dichotomized as “good” (“excellent” or “good”) or “poor” (“fair” or “poor”) (Wuorela et al., 2020). Happiness was assessed using the question, “How happy are you now?” with 11 answering options from 0 (very unhappy) to 10 (very happy), and the scores were dichotomized as “low” or “high” according to the cut-off point of 8 or higher which was shown in the previous studies (Ide et al., 2022; Moriyama et al., 2018). Depressive symptoms were assessed using the 15-item Geriatric Depression Scale (GDS) which was developed for self-administration in the community using a simple binary (yes/no) format (Wada et al., 2004). Participants were dichotomized as “not depressed” or “depressed” according to the cut-off point of five or higher for the GDS scores, which indicates mild to severe depression (Schreiner et al., 2003). Functional health was assessed using a subscale of the Tokyo Metropolitan Institute of Gerontology Index of Competence (Koyano et al., 1991). Among the five items, including travel, shopping, preparing meals, paying, and taking out and withdrawing savings, those with at least one difficulty were defined as “deficits” while others were “not deficits,” based on prior research (Fujihara et al., 2019). Community-level health and well-being assessment scores were defined by calculating their prevalences by aggregating them on a school district basis.

### Covariates

The community-level and individual-level sociodemographic characteristics were used as covariates in this study. As community-level variables, population density, aging proportion, and low education proportion were evaluated from national population census data, and each variable was divided into quartiles. As individual-level variables, we used age, gender, living arrangement, marital status, educational attainment, equivalent household income, and comorbidities. Age (years) was categorized as “65-69,” “70-74,” “75-79,” “80-84,” and “85 or older.” Living arrangement was dichotomized as “living with others,” and “living alone.” Marital status was categorized as “married,” “widowed/divorced,” “never married,” and “other.” Educational attainment (years) was categorized as “< 9”, “10-12,” and “≥ 13.” Equivalent household income was calculated by dividing the income of each household by the square root of the household size (number of family members), and categorized as “low” (< 2.00 million JPY), “middle” (2.00–3.00 million JPY), and “high” (≥ 3.00 million JPY). Comorbidities were categorized as “none,” “one,” “two,” and “three or more,” from a list of 17 illnesses, such as cancer, heart disease, stroke, diabetes, digestive diseases, dementia, and depression disorders.

### Statistical analysis

First, the descriptive statistics of the individuals and communities were calculated. Second, to assess the ecological relationships between the community-level AFC scores and the community-level health and well-being outcomes, we conducted partial correlations controlling for community-level covariates of population density, aging proportion, and low education proportion. In the analysis, all variables were scored at the ten percentile point and the Spearman’s rank correlation coefficient was calculated. Third, to examine the relationships between community-level AFC scores and individual health and well-being outcomes, we applied a multilevel Poisson regression analysis with level one as community and level two as individual, adjusted for all the individual- and community-level covariates and estimated the prevalence ratios (PRs) and 95% confidence intervals (CIs) for individual health and well-being. For the analysis, the three domain scores of the AFC scale were divided into quartiles on the community levels. To mitigate potential bias to missing information, we conduct missing-value imputation using chained random forests, based on the random forest algorithm (Stekhoven and Bühlmann, 2012).

The significance level was set at < 0.05. We used the R software (Version 4.3.1 for windows: R Foundation for Statistical Computing, Vienna, Austria) for all statistical analysis.

## Results

Data from 71,824 individuals and 145 communities where they lived were analyzed. Table 1 presents the characteristics of the participants. The mean age of the participants was 73.7 years (standard deviation = 6.2), and 53.9% were women. Among the participants, 12.6% showed poor self-reported health, 49.1% indicated a high level of happiness, 16.8% expressed depressive symptoms, and 43.3% had functional health deficits.

**Table 1.** The characteristics of the individuals.

|  |  | Overall |
| --- | --- | --- |
|  |  | n = 71824 |
| Age (years), n (%) | 65–69 | 23297 ( 32.4) |
|  | 70–74 | 19405 ( 27.0) |
|  | 75–79 | 15787 ( 22.0) |
|  | 80–84 | 9012 ( 12.5) |
|  | ≥ 85 | 4323 ( 6.0) |
| Gender, n (%) | Men | 33102 ( 46.1) |
|  | Women | 38722 ( 53.9) |
| Living arrangement, n (%) | Living with others | 58308 ( 81.2) |
|  | Living alone | 9130 ( 12.7) |
|  | Missing | 4386 ( 6.1) |
| Marital status, n (%) | Married | 52484 ( 73.1) |
|  | Widowed/divorced | 16287 ( 22.7) |
|  | Never married | 1572 ( 2.2) |
|  | Other | 475 ( 0.7) |
|  | Missing | 1006 ( 1.4) |
| Educational attainment (years), n (%) | < 10 | 27327 ( 38.0) |
|  | 10–12 | 28760 ( 40.0) |
|  | ≥ 13 | 15105 ( 21.0) |
|  | Missing | 632 ( 0.9) |
| Equivalent household income, n (%) | Low | 27285 ( 38.0) |
|  | Middle | 21039 ( 29.3) |
|  | High | 5795 ( 8.1) |
|  | Missing | 17705 ( 24.7) |
| Comorbidities, n (%) | None | 18554 ( 25.8) |
|  | One | 27884 ( 38.8) |
|  | Two | 15580 ( 21.7) |
|  | Three or more | 7397 ( 10.3) |
|  | Missing | 2409 ( 3.4) |
| Self-reported health, n (%) | Good | 62175 ( 86.6) |
|  | Poor | 9081 ( 12.6) |
|  | Missing | 568 ( 0.8) |
| Happiness, n (%) | Low | 34173 ( 47.6) |
|  | High | 35245 ( 49.1) |
|  | Missing | 2406 ( 3.3) |
| Depressive symptoms, n (%) | Not depressed | 47493 ( 66.1) |
|  | Depressed | 12071 ( 16.8) |
|  | Missing | 12260 ( 17.1) |
| Functional health, n (%) | Not deficits | 38708 ( 53.9) |
|  | Deficits | 31075 ( 43.3) |
|  | Missing | 2041 ( 2.8) |

Table 2 demonstrates the descriptive statistics for the community-level AFC scale. Regarding the domain of age-friendly physical environments, there were differences of approximately 40–50 points or more across communities, and the accessibility of transportation stops varied by approximately 70 points across communities. For the domain of social engagement and communication, community differences of approximately 15–35 points were found, but the difference in the item on internet use was the highest at 60 points. Regarding the domain of social inclusion and dementia-friendliness, although the item of friendship was small with a community difference of approximately 25 points, other items of social inclusion and respect for older adults and dementia-friendliness differed by more than approximately 30 points.

**Table 2.** Community-level descriptive statistics of the AFC scale (n = 145)

|  | Mean (SD) | Min–max |
| --- | --- | --- |
| **Age-friendly physical environments** |  |  |
| Accessibility of barrier-free public spaces and buildings | 14.0 (7.3) | 2.6–49.1 |
| Accessibility of barrier-free streets | 12.0 (7.0) | 0.0–37.0 |
| Outdoor space suitable for exercise | 73.0 (11.5) | 45.0–95.4 |
| Accessibility of barrier-free public transportation vehicles | 22.4 (8.6) | 2.6–44.4 |
| Accessibility of public transportation stops | 26.2 (14.0) | 1.2–70.7 |
| **Social engagement and communication** |  |  |
| Hobby group participation | 36.2 (6.0) | 19.8–54.1 |
| Sports group participation | 28.7 (6.3) | 13.3–43.8 |
| Learning and culture group participation | 8.5 (2.6) | 2.8–17.1 |
| Volunteer group participation | 14.9 (3.1) | 5.7–22.5 |
| Internet use | 42.8 (10.4) | 18.5–80.6 |
| **Social inclusion and dementia-friendliness** |  |  |
| Community belonging | 38.1 (11.1) | 16.1–79.2 |
| Involvement of community decision | 43.2 (11.6) | 18.6–78.8 |
| Perception of community reciprocity | 55.0 (5.8) | 44.1–70.8 |
| Friendships | 74.8 (4.2) | 60.0–85.4 |
| Community support and health services | 45.7 (9.0) | 20.3–70.0 |
| Social participation of people with dementia | 50.9 (7.2) | 34.6–71.0 |
| Support for families of people with dementia | 78.7 (6.2) | 61.1–94.3 |

AFC, age-friendly community; SD, standard deviation.
All items were scored from 0 to 100.

Table 3 shows the results of the ecological correlations between community-level AFC scale scores and community-level health and well-being prevalences. After adjusting for community-level covariates, higher age-friendly physical environments were inversely correlated with functional health deficits (r = −0.412, *P* < 0.001). Higher social engagement and communication were inversely correlated with poor self-reported health (r = −0.216, *P* = 0.010), depressive symptoms (r = −0.424, *P* < 0.001) and functional health deficits (r = −0.277, *P* < 0.001). Social inclusion and dementia-friendliness were inversely correlated with poor self-reported health (r = −0.223, *P* = 0.008) and depressive symptoms (r = −0.247, *P* = 0.003), and positively correlated with higher levels of happiness (r = 0.247, *P* < 0.001).

**Table 3.** Community-level correlations of the AFC scale with health and well-being (n = 145)

|  | Partial correlation coefficients† |  |  |  |
| --- | --- | --- | --- | --- |
|  | Poor self-reported health | Higher levels of happiness | Depressive symptoms | Functional health deficits |
| *AFC scale* |  |  |  |  |
| Age-friendly physical environments | -0.087 | 0.054 | -0.129 | -0.412*** |
| Social engagement and communication | -0.216** | 0.189 | -0.424*** | -0.277*** |
| Social inclusion and dementia-friendliness | -0.223** | 0.276*** | -0.247** | 0.126 |

\*\*, $P < 0.01$ ; \*\*\*, $P < 0.001$ . AFC, age-friendly community.
†Spearman's rank correlation coefficients adjusted for population density, older adult proportion, and low education proportion at the community level.

Table 4 exhibits the association between community-level AFC scale scores and individual-level outcomes, based on multilevel Poisson regression analysis. Regarding self-reported health, neither the domains of the AFC showed significant associations. For happiness, the domain of social inclusion and dementia-friendliness was positively associated with its higher levels (per 25th percentile points: PR = 1.01 [95% CI = 1.001–1.02], *P* = 0.028). Regarding depressive symptoms, the domain of social engagement and communication were inversely associated with the individual likelihood of this condition (per 25th percentile points: PR = 0.96 [95% CI = 0.93–0.98], *P* < 0.001). For functional health, the domain of age-friendly physical environments was inversely associated with individual functional health deficits (per 25th percentile points: PR = 0.98 [95% CI = 0.97–0.995], *P* = 0.006).

**Table 4.**
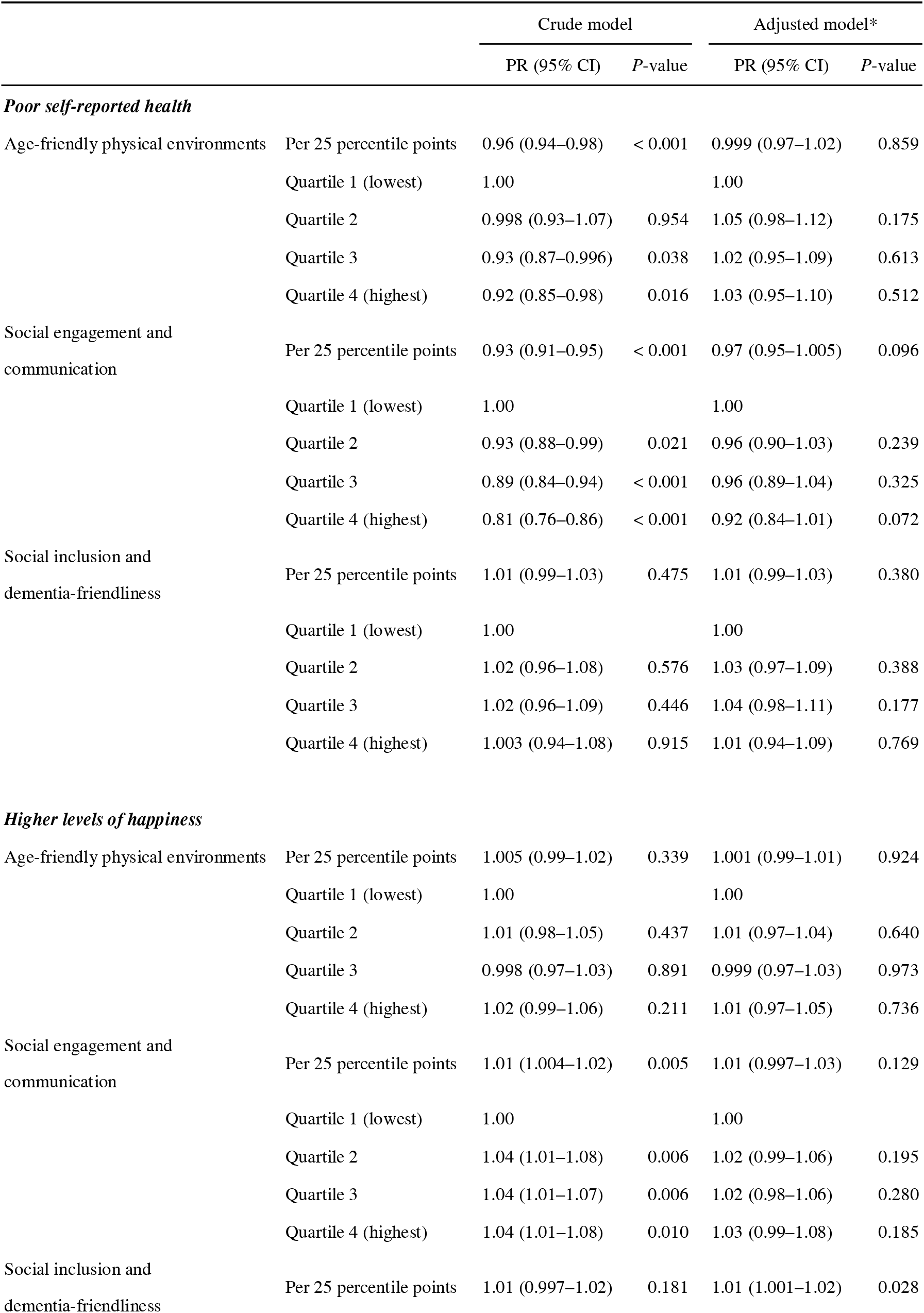
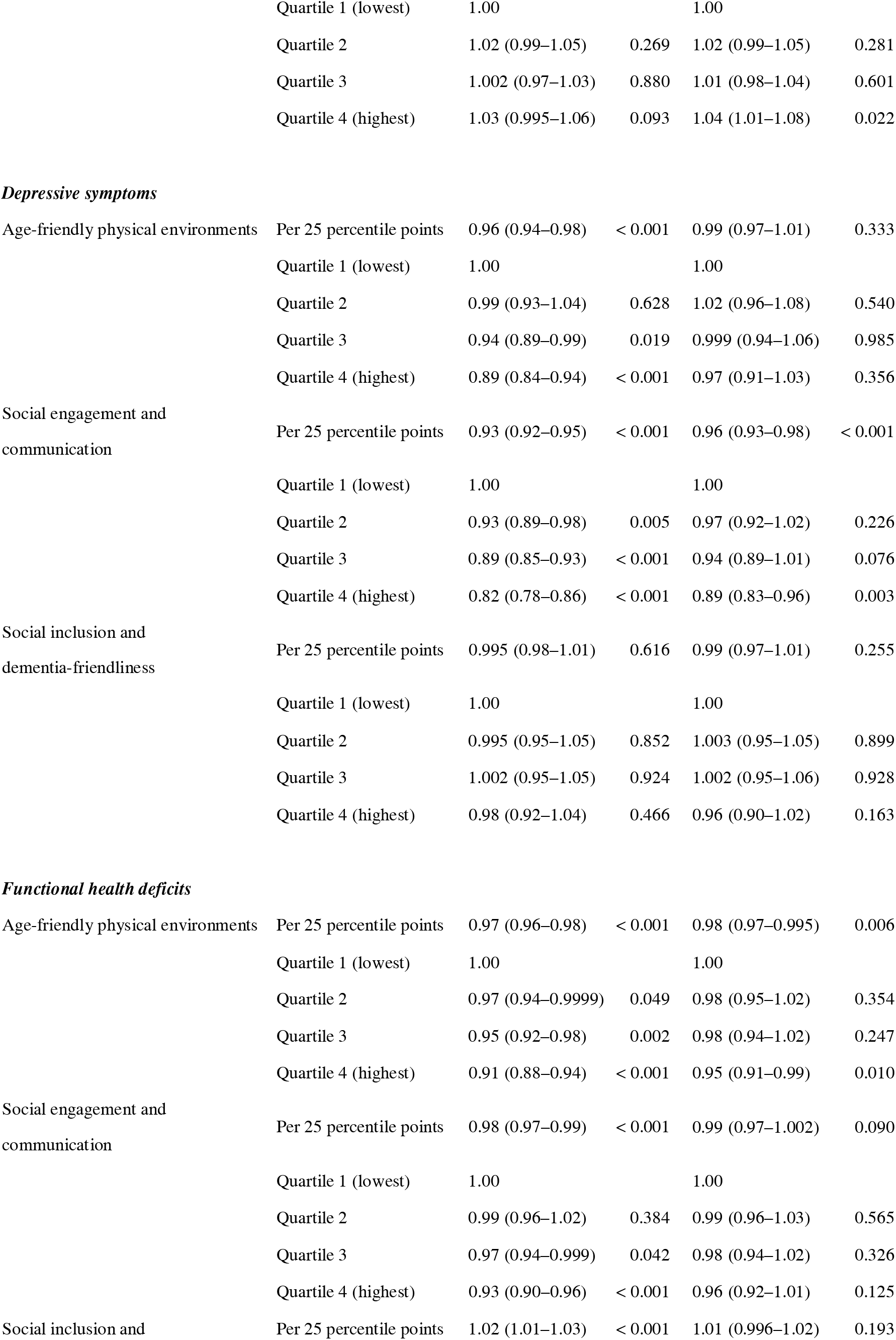
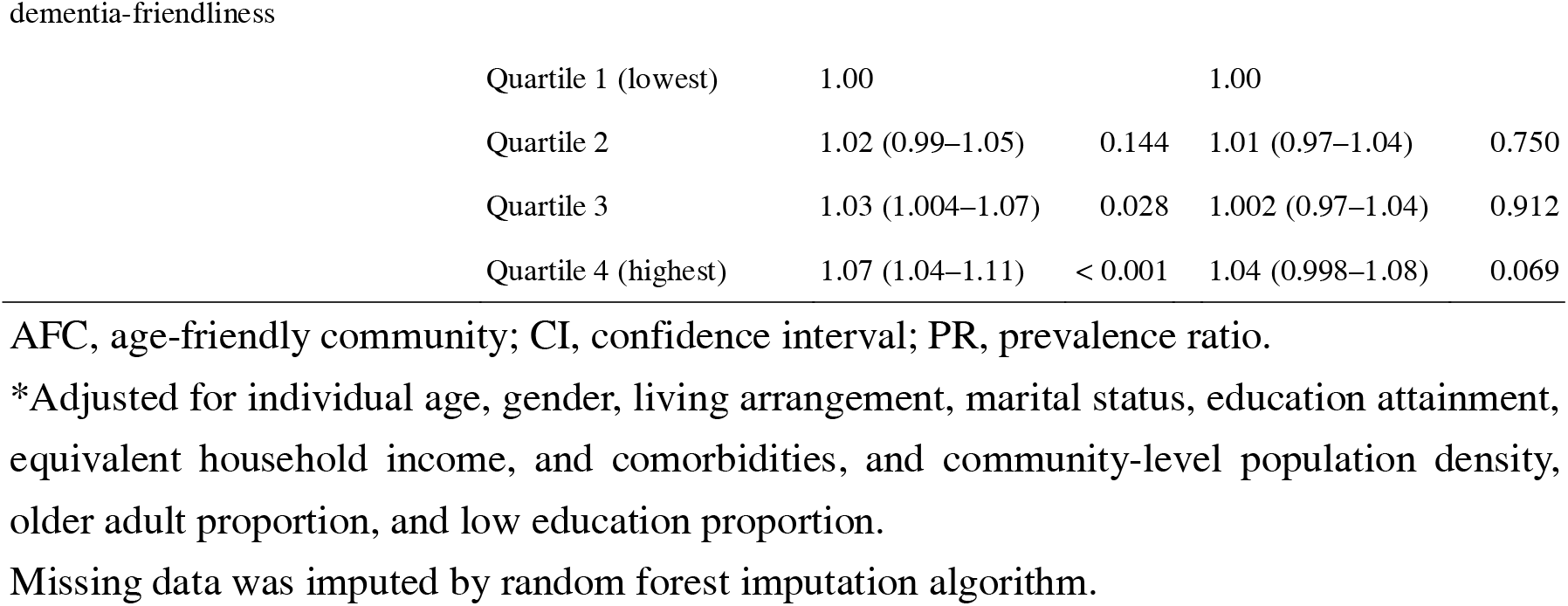
Association of the AFC scale scores with individual-level health and well-being, based on multilevel Poisson regression analysis (n = 71,824)

Table 5 provides a summary of the results of the ecological and multilevel analyses. Across both analyses, the age-friendly physical environment domain was consistently associated with functional health, the social engagement and communication domain showed consistent associations with depressive symptoms and functional health, and finally, the social inclusion and dementia-friendliness domain consistently showed links to a sense of happiness.

**Table 5.** Summary of the results of the ecological and multilevel analyses.

|  |  | Health and well-being outcomes |  |  |  |
| --- | --- | --- | --- | --- | --- |
|  |  | Self-reported<br>health | Happiness | Depressive<br>symptoms | Functional<br>health |
| Age-friendly physical environments | Ecological analysis |  |  |  | X |
|  | Multilevel analysis |  |  |  | X |
| Social participation and communication | Ecological analysis | X |  | X | X |
|  | Multilevel analysis |  |  | X |  |
| Social inclusion and dementia-friendliness | Ecological analysis | X | X | X |  |
|  | Multilevel analysis |  | X |  |  |

X indicates that a statistically significant association was found.

## Discussion

This ecological and multilevel study examined the association of community-level age-friendliness with health and well-being. Through both analyses, the AFC consistently linked each of the three domains to the health and well-being indicators of older adults. The domain of age-friendly physical environments reflected functional health, the domain of social engagement and communication was reflective of depressive symptoms, and social inclusion and dementia-friendliness reflected happiness. This means that community-level age-friendliness, along with the three domains, provides a well-reflection of the overall health and well-being of older adults. Our findings suggest the importance of implementing the AFC through community evaluations and related policy promotions.

Each of the three AFC domains in this study reflected different outcome measures, supported by the fact that various features of the AFC are associated with different outcomes (Meeks, 2022). The domain of age-friendly physical environments involves the enrichment of outdoor spaces and transportation resources accessible to older adults. Such an environment has been shown to contribute to maintaining well higher-order living activities of older adults, which includes shopping and monetary management (Pan et al., 2024). This means that community physical environments that make activities of daily living more accessible to older adults may provide assistance for their functional health. Meanwhile, the domain of social engagement and communication consists of the social activity and employment of older adults in the community and accessibility to information and communication. Prior research has reported that older adults are less depressed in communities with higher social participation (Yamaguchi et al., 2019). Additionally, access to information and communication, such as internet use, positively affects older adults’ social participation (Kondo et al., 2021; Nakagomi et al., 2022), which may improve their mental health. These support the notion that communities in which older adults are active may contribute well to residents’ mental health. In the other, the domain of social inclusion and dementia-friendliness encompasses the inclusiveness of older adults in the community, including living well with individuals with dementia. Communities with supportive and inclusive relations have been shown to be associated with higher levels of well-being and positive mental health among residents, suggesting the contributions to their happiness and well-being (Cramm and Nieboer, 2015; Cramm et al., 2013); these were also in line with the previous finding that the built environment is less important to psychosocial health (Kim et al., 2022). Given that health and well-being comprise multiple aspects, promoting comprehensive fields of the AFC may help the health and well-being of older adults.

In this study, the ecological analysis showed that each AFC domain was multiply associated with health and well-being outcomes. Particularly, the domains of social engagement and communication and social inclusion and dementia-friendliness reflected well the positive results of several of the indicators. These results highlight that AFC mirrors the health and well-being of older residents. Given that, in the process of building AFC, it is crucial to measure, evaluate, and monitor the age-friendliness of the community (Dikken et al., 2020), our findings support the meaning of evaluating it using the measurement of the multi-dimensions of AFC well reflected the health and well-being (Meeks, 2022); it encourages policymakers to monitor the AFC status in the healthy aging policy process.

The present study used a multidimensional scale that covered the AFC core areas described by the WHO, along with an additional element of dementia-friendliness. In communities with respect and inclusion for older adults, including individuals living with dementia, the residents had a higher level of sense of happiness. The WHO states that the AFC should be promoted in consideration of or complemented by the DFC (World Health Organization, 2021). Some initiatives have been made to adopt the AFC toward the DFC (Turner and Cannon, 2018). These policy trends underscore the importance of community evaluation and monitoring of AFC scales that take into account dementia-friendliness. Our findings update the AFC framework through assessing some dementia-friendliness elements. Because this study only covered part of the dementia-friendliness, further research is needed for capturing comprehensive the DFC. Nevertheless, we believe that this study may help policymakers and public health practitioners in community development to achieve healthy aging and aging-in-place, including individuals with dementia.

This study holds significance as the first in terms of examining the association of community-level age-friendliness with individual health and well-being, using the assessment scale considering dementia-friendliness aspects, through ecological and multilevel analyses. However, there are several study limitations. First, the nature of the cross-sectional analysis could not address the causality, requiring attention to reverse causality. This means that depending on their health and well-being conditions, older adults may reside in more age-friendly areas. Further investigations with longitudinal data are needed to test the long-term impact of health and well-being outcomes. Second, the data were based on community-dwelling older adults without public long-term care insurance benefits. Therefore, the results were derived from data from relatively healthy older adults, which may not necessarily fully reflect the impact of including older adults with disabilities or dementia within the community. Future research should evaluate the relationships with the health and well-being of older adults with care needs and cognitive impairment. Third, the AFC scale in this study covered only aspects of the social environment, such as the supportiveness and inclusiveness of people with dementia and their families, among the dementia-friendliness elements. Hence, this scale fails to appreciate aspects of the physical environments related to dementia-friendliness, such as signs and design in the community (Dementia Australia; Turner and Cannon, 2018). Additional examinations using these updated scales are necessary. Finally, this study used data from a wide range of urban to rural areas in Japan, but the number of communities covered in the final analysis was limited to 145 to ensure the precision of the measurement in the respective areas, which may limit the representativeness of the entire area in Japan. Future studies using data from nationwide community samples are required to test the generalizability of this scale.

## Conclusions

This study revealed that community-level age-friendliness, including elements of dementia-friendliness, was associated with health and well-being among older adults, by ecological and multilevel analysis. Our findings help develop communities that promote healthy aging of older adults including individuals living with dementia, and their social inclusion, through facilitating AFC.

## Data Availability Statement

For the JAGES, all inquiries are to be addressed to the data management committee via. All JAGES datasets have ethical or legal restrictions for public deposition because of the inclusion of sensitive information about the human participants.

## Acknowledgments

We wish to express our sincere gratitude to the staff in the surveyed municipalities for their contributions. We also thank the members of the 300 BM and AFC working group of the Japan Gerontological Evaluation Study.

## Author Contributions

TN conceptualized and designed the study, analyzed the data, interpreted the results, and drafted and revised the manuscript. SF conceptualized and designed the study, analyzed the data, help to interpret the results, and reviewed and critically revised the manuscript. KI, SJ and TS helped to develop the study concept, and reviewed and critically revised the manuscript. KK, the principal investigator of the JAGES, helped to develop the study concept, participated in data collection and study design, and reviewed and critically revised the manuscript. TO, the project leader, participated in data collection, helped to develop the study concept, and interpret the results, and reviewed and critically revised the manuscript.

