## supplementary for "Association of age-friendly communities with health and well-being among older adults: an ecological and multilevel analysis from the Japan Gerontological Evaluation Study"

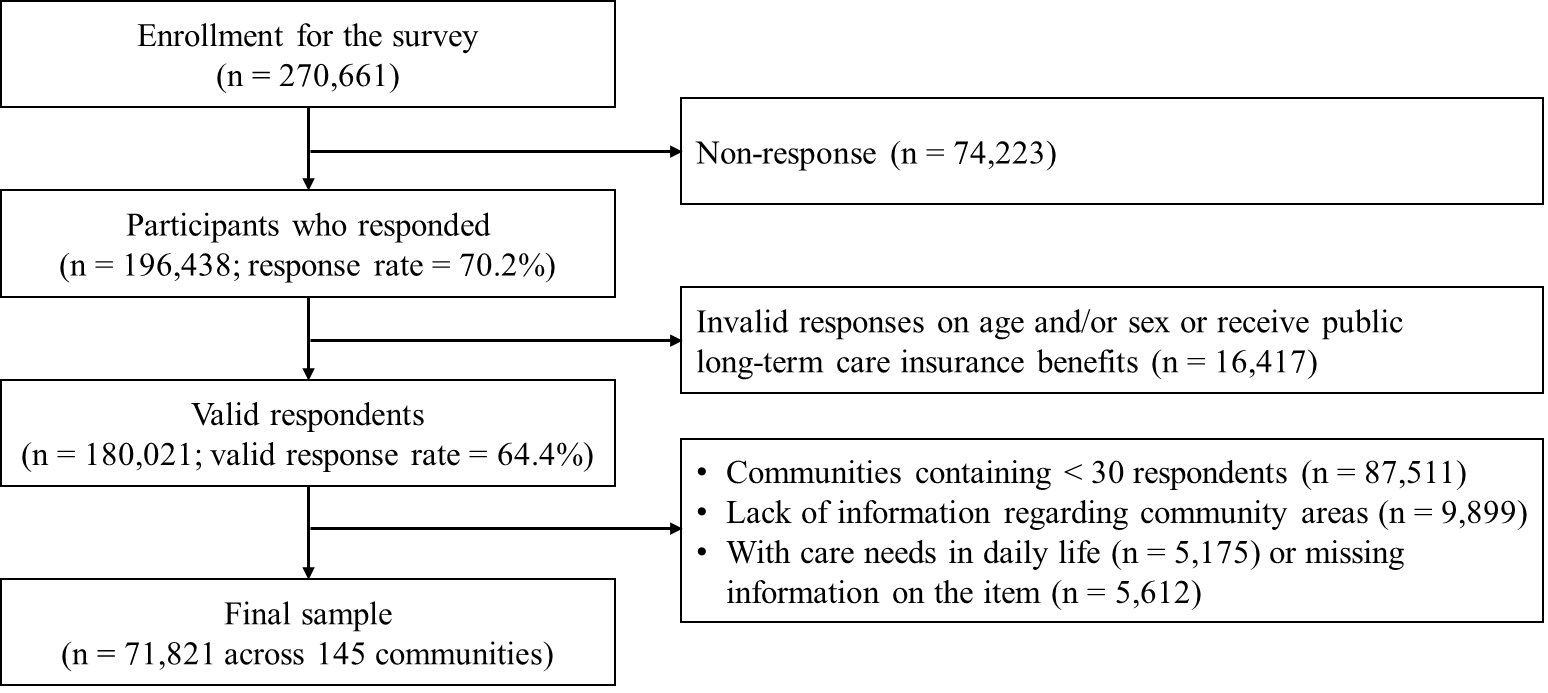

**Supplementary Figure 1. Sample selection flow chart**

**Supplementary Table 1. Items regarding the age-friendly community scale**

| AFC domains | Items | Questions and responses |
| --- | --- | --- |
| **Age-friendly physical environments** |  |  |
| Outdoor spaces and buildings | Accessibility of barrier-free public spaces and buildings | Do you have any public facilities available for individuals experiencing difficulty walking or visual/hearing disabilities in your area? (responses: “a lot,” “somewhat,” “not much,” “not at all,” or “unknown”; 1 = “a lot” or “somewhat,” 0 = others) |
| Transportation | Accessibility of barrier-free streets | Do you have any streets available for individuals with wheelchairs, canes, or walkers to walk on in your neighborhood? (responses: “a lot,” “somewhat,” “not much,” “not at all,” or “unknown”; 1 = “a lot” or “somewhat,” 0 = others) |
| Outdoor spaces and buildings | Outdoor space suitable for exercise | Do you have any parks or sidewalks suitable for exercise or walking in your area? (responses: “a lot,” “somewhat,” “not much,” “not at all,” or “unknown"; 1 = "a lot" or "somewhat," 0 = others) |
| Transportation | Accessibility of barrier-free public transportation vehicles | Do you have any trains or buses available for individuals experiencing difficulty walking or visual/hearing disabilities in your area? (responses: “a lot,” “somewhat,” “not much,” “not at all,” or “unknown”; 1 = “a lot” or “somewhat,” 0 = others) |
| Transportation | Accessibility of public transportation stops | Do you have any train or subway stations in your neighborhood? (responses: “a lot,” “somewhat,” “not much,” “not at all,” or “unknown”; 1 = “a lot” or “somewhat,” 0 = others) |
| **Social engagement and communication** |  |  |
| Social participation | Participation in hobby groups | How often do you participate in hobby groups? (responses: “4 times or more a week,” “twice to three times a week,” “once a week,” “once to three times a month,” “sometimes a year,” or “none”; 1 = once or more a month, 0 = others) |
| Social participation | Participation in sports group and club | How often do you participate in sports group and club? (responses: “4 times or more a week,” “twice to three times a week,” “once a week,” “once to three times a month,” “sometimes a year,” or “none”; 1 = once or more a month, 0 = others) |
| Social participation | Participation in learning and cultural activities | How often do you participate in learning and cultural activities? (responses: “4 times or more a week,” “twice to three times a week,” “once a week,” “once to three times a month,” “sometimes a year,” or “none”: 1 = once or more a month, 0 = others) |
| Civic participation and employment | Participation in volunteer groups | How often do you participate in volunteer groups? (responses: “4 times or more a week,” “twice to three times a week,” “once a week,” “once to three times a month,” “sometimes a year,” or “none”; 1 = once or more a month, 0 = others) |
| Communication and information | Internet use | Have you used the Internet or e-mail in the past year? (responses: “none,” “sometimes a month,” “twice to three times a week,” or “almost everyday”; 1 = “sometimes a month” to “almost everyday,” 0 = “none”) |
| **Social inclusion and dementia-friendliness** | |  |
| Respect and social inclusion | Sence of belonging | Do you think you're respected by your neighbors and that you're a member of your community? (responses: “agree,” “somewhat agree,” “either,” “somewhat disagree,” or “disagree”; 1 = “agree” or “somewhat agree” or 0 = others) |
| Respect and social inclusion | Participation in community decisions | Do you participate in decision-making in your community by attending your neighborhood association or similar community meetings? (responses: “agree,” “somewhat agree,” “either,” “somewhat disagree,” or “disagree”; 1 = “agree” or “somewhat agree,” 0 = others) |
| Respect and social inclusion | Perception of community reciprocity | Do you think individuals living in your area try to help others in the most of situations? (responses: “agree,” “somewhat agree,” “either,” “somewhat disagree,” or “disagree”; 1 = “agree” or “somewhat agree,” 0 = others) |
| Communication and information | Frequency of meeting with friends | How often do you meet friends? (responses: “almost everyday,” “twice to three times a week,” “once a week,” “once to three times a month,” sometimes a year,” “none”; 1 = “once or more a month,” 0 = others) |
| Community support and health services | Community health care service | Do the local government offices and private companies in your area by and large offer welfare services necessary for your daily life and health? (responses: “agree,” “somewhat agree,” “either,” “somewhat disagree,” or “disagree”; 1 = “agree” or “somewhat agree,” 0 = others) |
| Dementia-friendliness | Social participation of people with dementia | Do you think individuals with dementia should take part in community activities and have some role in those activities? (responses: “agree,” “somewhat agree,” “either,” “somewhat disagree,” or “disagree”; 1 = “agree” or “somewhat agree,” 0 = others) |
| Dementia-friendliness | Support for families of people with dementia | If one of your family members were affected by dementia, would you like your neighbors and acquaintances to know about it so that you could get assistance from them? (responses: “agree,” “somewhat agree,” “either,” “somewhat disagree,” or “disagree”; 1 = “agree” or “somewhat agree,” 0 = others) |

AFC, age-friendly community.

**Supplementary Table 2. Factor loadings of the age-friendly community scale**

|  | Factor 1 (Age-friendly physical environments) | Factor 2 (Social engagement and communication) | Factor 3 (Social inclusion and dementia-friendliness) | Communality | Unique variance | Complexity |
| --- | --- | --- | --- | --- | --- | --- |
| Accessibility of public transportation vehicles | **0.849** | -0.042 | 0.109 | 0.63 | 0.37 | 1.04 |
| Accessibility of public transportation stops | **0.760** | -0.039 | -0.080 | 0.61 | 0.39 | 1.03 |
| Accessibility of unimpeded streets | **0.730** | -0.036 | -0.024 | 0.52 | 0.48 | 1.01 |
| Accessibility of public spaces and buildings | **0.719** | 0.033 | 0.121 | 0.49 | 0.51 | 1.06 |
| Outdoor space suitable for exercise | **0.605** | 0.236 | 0.008 | 0.55 | 0.45 | 1.30 |
| Hobby group participation | -0.026 | **0.985** | -0.074 | 0.96 | 0.04 | 1.01 |
| Sports group participation | 0.001 | **0.915** | -0.033 | 0.84 | 0.16 | 1.00 |
| Volunteer group participation | -0.123 | **0.677** | 0.151 | 0.42 | 0.58 | 1.17 |
| Learning and culture group participation | 0.233 | **0.607** | 0.070 | 0.54 | 0.46 | 1.32 |
| Internet use | 0.215 | **0.418** | -0.259 | 0.43 | 0.57 | 2.23 |
| Community belonging | -0.110 | -0.079 | **0.783** | 0.72 | 0.29 | 1.06 |
| Perception of community reciprocity | -0.120 | 0.115 | **0.754** | 0.64 | 0.36 | 1.10 |
| Community support and health services | 0.092 | 0.091 | **0.743** | 0.51 | 0.49 | 1.06 |
| Involvement of community decision | -0.271 | -0.013 | **0.696** | 0.71 | 0.29 | 1.30 |
| Positive attitude toward social participation of people with dementia | 0.259 | 0.081 | **0.578** | 0.30 | 0.70 | 1.43 |
| Help-seeking as a family member of people with dementia | 0.124 | -0.252 | **0.574** | 0.34 | 0.66 | 1.48 |
| Friendships | -0.416 | 0.202 | **0.450** | 0.47 | 0.53 | 2.39 |
| Correlation coefficients between factors |  |  |  |  |  |  |
| Factor 1 | 1.000 | 0.475 | -0.396 |  |  |  |
| Factor 2 |  | 1.000 | -0.063 |  |  |  |
| Factor 3 |  |  | 1.000 |  |  |  |

Exploratory factor analysis was applied Promax rotation and maximum likelihood method.
